# Shared Data As a Mechanism for Integration between Housing and Health Sectors: A Qualitative Interview Study in the UK

**DOI:** 10.64898/2026.10.20.26366050

**Authors:** Oluwadolapo Adegboye, Eleanor Wyke, Sana Ali, Jessica Veltman, Jessica Sheringham

## Abstract

**Background:** Data sharing is widely positioned as a key mechanism for enabling integration between health and non-health sectors, particularly in relation to wider determinants such as housing. However, there is limited empirical understanding of how cross-sector data sharing is enacted in practice.

**Methods:** Semi-structured interviews were undertaken with professionals from housing, NHS, public health and voluntary and community sector organisations in the UK. Data were analysed using reflexive thematic analysis, informed by Normalisation Process Theory. A typology of data sharing models was developed inductively.

**Results:** From 27 interviews, six models were identified through which data sharing enabled and sustained integration across sectors. Models ranged from referral pathways supporting individual-level coordination to linked datasets supporting population-level service planning and monitoring. Participants balanced potential benefits against concerns relating to privacy, trust and organisational capacity. While initiatives supported improved coordination and greater recognition of housing as a determinant of health, some remained emergent and unevenly embedded.

**Discussion:** Integration depended not only on digital infrastructure, but on the relational and organisational work required to legitimise and operationalise data sharing.

**Conclusion:** Policies emphasising digital infrastructure as a means to integration should be complemented by attention to the relational, organisational and ethical conditions required to support effective cross-sector data sharing.

## Introduction

Data or information sharing is widely recognised as a core enabler to integration within healthcare [1]. Data sharing, encompassing both communication between practitioners and joined up information systems, can facilitate monitoring of process and outcomes across boundaries and its absence can be a barrier to achieving the desired outcomes of integration, e.g. continuity of care.

There is increasing recognition of the value of bringing together data across sectors, specifically across housing and health, to better understand the relationships between place, housing conditions, and health outcomes, particularly for population health research and service planning [2,3]. There is growing evidence that integration between housing and health sectors could lead to better outcomes [4]. In England, for example, poor housing has multiple and severe impacts on health, costing the NHS in England over £1.4 billion annually [5]. Siloed working means those across housing and health services often lack access to the information needed to understand residents’ holistic needs, limiting opportunities for coordinated responses [6]. Recent policy developments, including an emerging policy priority for neighbourhood health, position cross-sector collaboration and data sharing as central to preventive, place-based approaches to improving population health [7].

It is well established that cross-sector data sharing is often challenging and uneven, with barriers including: unclear or interdependent governance frameworks and legislative concerns; lack of interoperable IT systems; limited training and skills; and organisational culture [1,8,9,10]. Yet, there remains an evidence gap on where and how data sharing between housing and health can progress [10,6,3]. For example, Tweed et al’s scoping reviews of data sharing identified mainly studies within healthcare. They found no studies of cross-sector health and housing research and studies were largely focused on data sharing for research purposes, rather than how cross-sector data sharing is enacted in frontline service contexts [10].

This study seeks to develop understanding of how data sharing between housing and health develops in service settings. It examines what motivates individuals within organisations to initiate data sharing, how professionals negotiate associated risks and benefits, and what such initiatives are perceived to achieve.

## Methods

### Design

This study used a qualitative interview design. A description of the planned methods is available in the research protocol [11] but key details and developments since the protocol was written are provided below. A qualitative approach was chosen to enable in-depth exploration of participants’ experiences, interpretations and practices relating to cross-sector data sharing and integration. Ethical approval was granted by the UCL Life and Medical Sciences Research Ethics Committee (Ethics reference: 0128). All participants provided informed written consent prior to taking part.

Normalisation Process Theory (NPT; [12]), was used throughout the study as a sensitising and organising framework to understand how data sharing functioned as a mechanism through which integration between housing and health was produced, negotiated and stabilised over time. NPT focuses on how new practices are implemented, embedded and sustained in routine work.

### Involvement of people with lived experience

The study is part of a wider programme of research on housing and health [11]. The design of this programme involved consultation with residents and resident representatives in one borough of London. This consultation informed the decision not to include residents within this study but to conduct separate research with them.

The consultation also informed the broad framing of “data sharing” taken in the study. Data sharing encompassed both formal and less formal mechanisms to share both individual or population-level information. Information encompassed not only data on resident or patient records but also qualitative insights gained from discussions with patients or residents. This broad framing was adopted because we recognised data sharing meant different things to different people and to prevent excluding relevant examples that interviewees may or may not have considered as “data sharing”. In particular, lived experience perspectives alerted us to the risk that adhering to a narrow definition may miss less formalised ways of data sharing.

### Setting and context

The study interviewed individuals across multiple regions in England, Wales and Scotland, involving partnerships between NHS, local authority (including public health teams), housing providers and voluntary, community, and social enterprise (VCSE) sector organisations.

The work was situated within the English policy context of Integrated Care Systems (ICSs), where data sharing between health and local government is positioned as a key mechanism for addressing wider determinants of health and reducing inequalities [13].

### Sampling and recruitment

A purposive sampling strategy [14] was used to capture a range of perspectives on data sharing across sectors and roles. Participants were invited to participate based on their involvement in, or experience of, data sharing initiatives between housing and health.

We sought a balance of individuals in strategic, operational and analytical roles, enabling exploration of both system-level and frontline perspectives. Participants were recruited through meetings (online or in person) or emails shared through professional networks, existing collaborations, and snowball sampling, including contacts within ICS partnerships, housing services and VCSE organisations, or through direct email invitations to specific individuals identified as potentially relevant by the study team. An information sheet with researcher details and the goals of the research was emailed to all prospective participants following an expression of interest in taking part.

Throughout the paper, “organisations” refers to the individual provider or commissioner entities - health, social housing provider and VCSE. “Sector” refers to broad purpose of organisations. “Partners” is used when describing collaborative relationships between organisations. “Residents”, “patients” or “clients” are used according to the sectoral context being described.

### Data collection

Data were collected through semi-structured interviews by interviewers trained in qualitative research methods (EW, OA, JS, JV) between January and November 2025. Less experienced interviewers conducted initial interviews jointly with JS initially, who also reviewed transcripts/recordings and any field notes made during the interview with interviewers. In one instance, one interviewee declined the request for audio recording, and their data were not included in the dataset.

An interview topic guide (see supplementary data) was piloted and then adapted for each subsequent interview. It first asked participants to describe specific examples of data sharing initiatives. Then, informed by Normalisation Process Theory, interviews explored:

- how participants understood the purpose and value of data sharing (coherence);
- how data sharing was introduced, supported and legitimised within organisations (cognitive participation);
- how participants evaluated the effects and consequences of data sharing (reflexive monitoring); and
- how data sharing was enacted in practice, including workflows, tools and responsibilities (collective action).

Interviews were conducted via Microsoft Teams (lasting 45-60 minutes), audio-recorded with consent, and transcribed verbatim by a professional transcription company, with transcripts checked by interviewers for accuracy.

### Data analysis

Data were analysed by EW, SA, OA and JS using a reflexive thematic approach [15], supported by NVivo software.

Analysis proceeded in three stages:

#### 1. Familiarisation and rapid appraisal

Each interview was summarised using a structured rapid appraisal template (supplementary data) to capture:

- context and actors involved;
- type and direction of data sharing;
- mechanisms and purposes of sharing; and
- perceived outcomes and impacts.

These summaries were collated into a database to enable comparison across cases (see Tables 1-2).

**Table 1.** Participant characteristics (n = 27)

| **Characteristic** | **Category** | **n** |
| --- | --- | --- |
| **Sector** | Housing / Local Authority* | 14 |
|  | NHS / Health | 10 |
|  | Voluntary, Community, and Social Enterprise Sector (VCSE) | 3 |
| **Role Level** | Strategic / Senior (Director, Lead, Consultant, Manager) | 15 |
|  | Other / Mixed (operational, specialist, technical roles) | 12 |
| **Geographic Coverage** | Yorkshire & Humber | 5 |
|  | London (North Central + North East) | 7 |
|  | East of England | 4 |
|  | South West | 2 |
|  | Scotland | 1 |
|  | East Midlands | 3 |
|  | West Midlands | 3 |
|  | National/ multi-region | 2 |

**Note:** \*Includes local authority housing and public health teams, housing associations, and related housing-sector organisations.

**Table 2.**
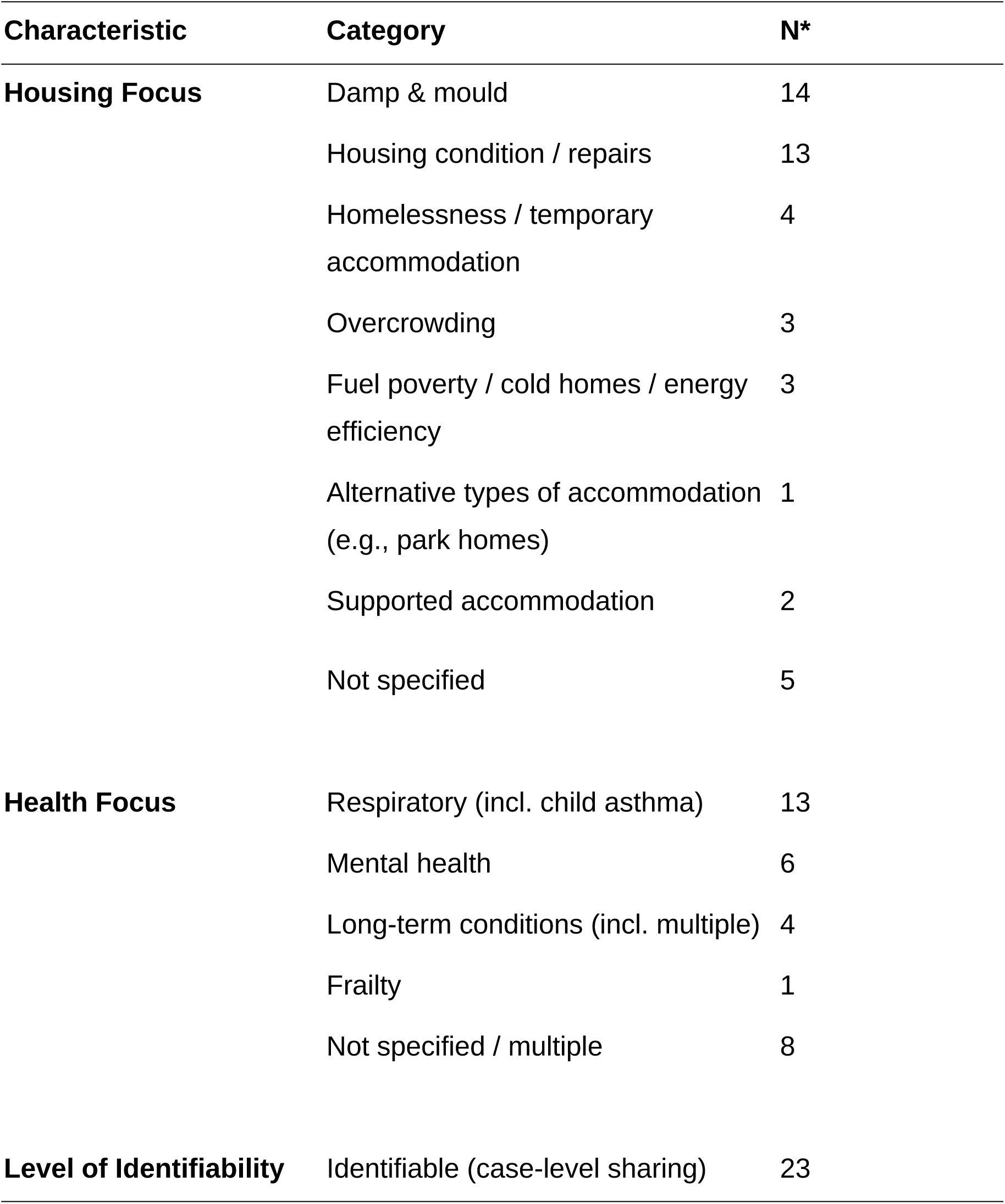
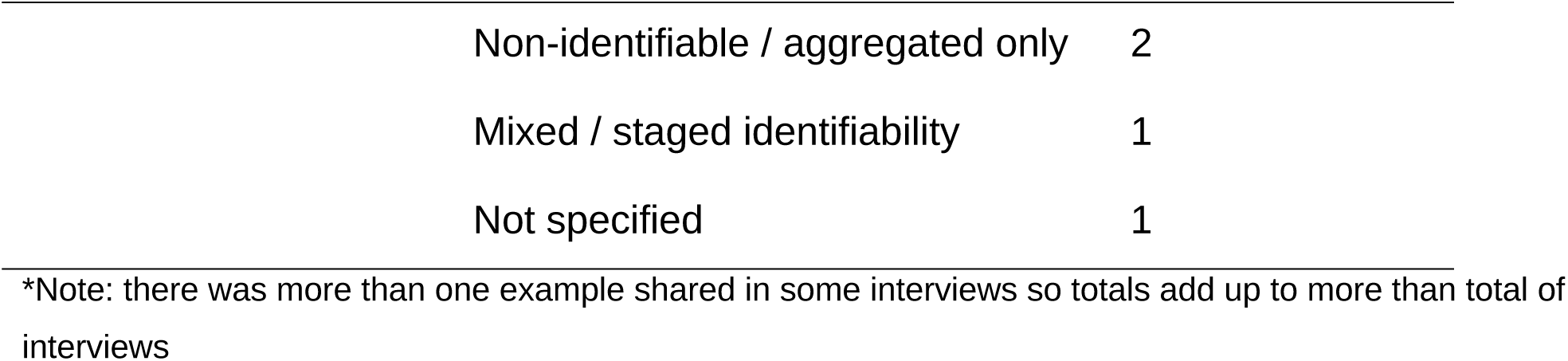
Characteristics of data sharing initiatives.

| **Characteristic** | **Category** | **N\*** |
| --- | --- | --- |
| **Housing Focus** | Damp & mould | 14 |
|  | Housing condition / repairs | 13 |
|  | Homelessness / temporary accommodation | 4 |
|  | Overcrowding | 3 |
|  | Fuel poverty / cold homes / energy efficiency | 3 |
|  | Alternative types of accommodation (e.g., park homes) | 1 |
|  | Supported accommodation | 2 |
|  | Not specified | 5 |
| **Health Focus** | Respiratory (incl. child asthma) | 13 |
|  | Mental health | 6 |
|  | Long-term conditions (incl. multiple) | 4 |
|  | Frailty | 1 |
|  | Not specified / multiple | 8 |
| **Level of Identifiability** | Identifiable (case-level sharing) | 23 |

|  |  |
| --- | --- |
| Non-identifiable / aggregated only | 2 |
| Mixed / staged identifiability | 1 |
| Not specified | 1 |

\*Note: there was more than one example shared in some interviews so totals add up to more than total of interviews

#### 2. Coding and theme development

Transcripts were coded inductively in NVivo, with NPT used as a sensitising framework (see supplementary data for codebook illustrating how NPT was interpreted for the study with examples of coded text). NPT constructs were used to inform interpretation rather than being imposed rigidly, allowing themes to develop through iterative interpretation of the data. All interviews were dual coded.

#### 3. Development of data sharing models

A typology of data sharing models was developed inductively from the interview data. These models captured distinct ways in which data were shared (e.g. referral-based, linked datasets), including:

- purpose;
- mechanisms of sharing;
- level of identifiability; and
- typical contexts of use.

This typology (Table 3) enabled comparison of how integration was produced through different configurations of data sharing practices.

**Table 3.**
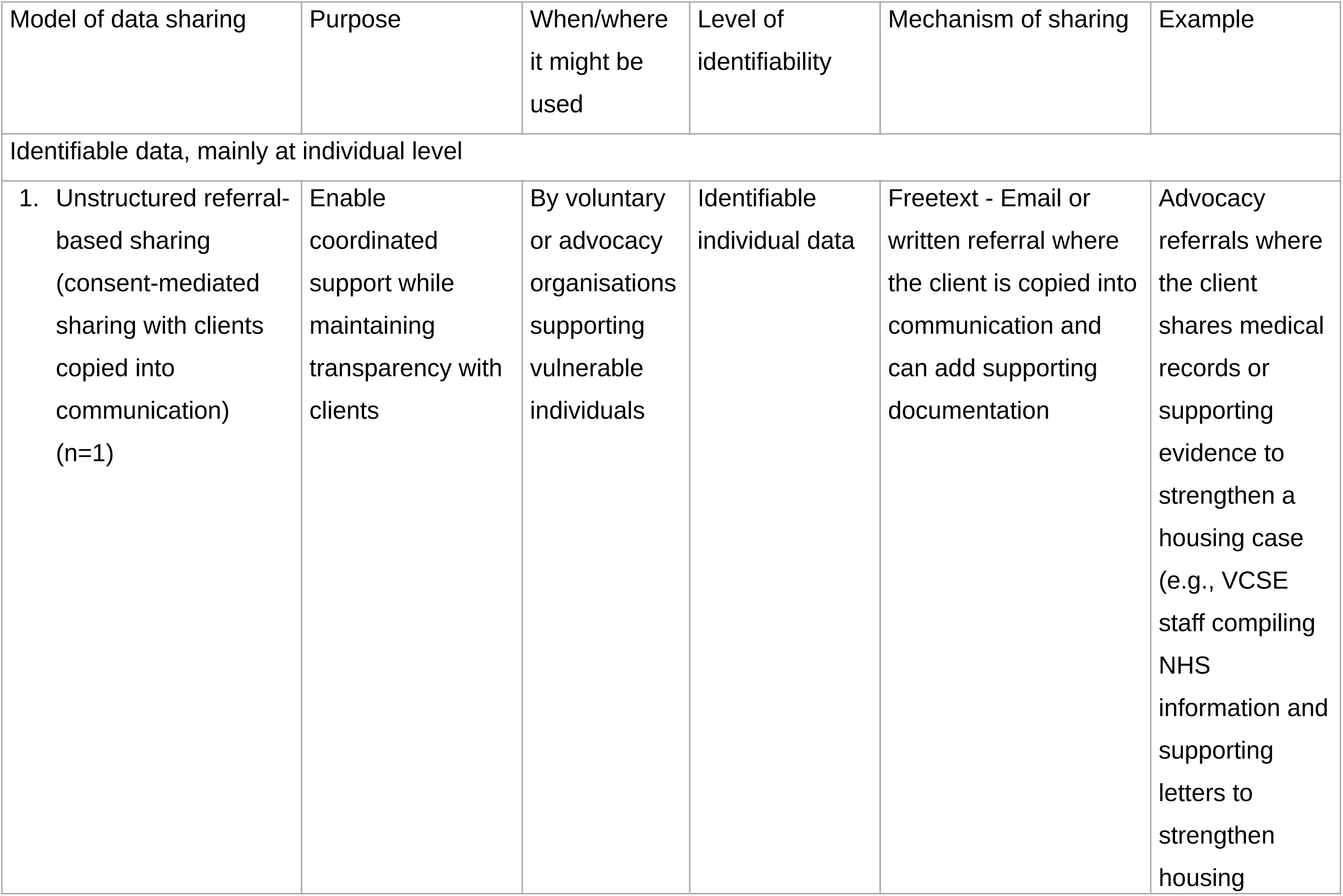
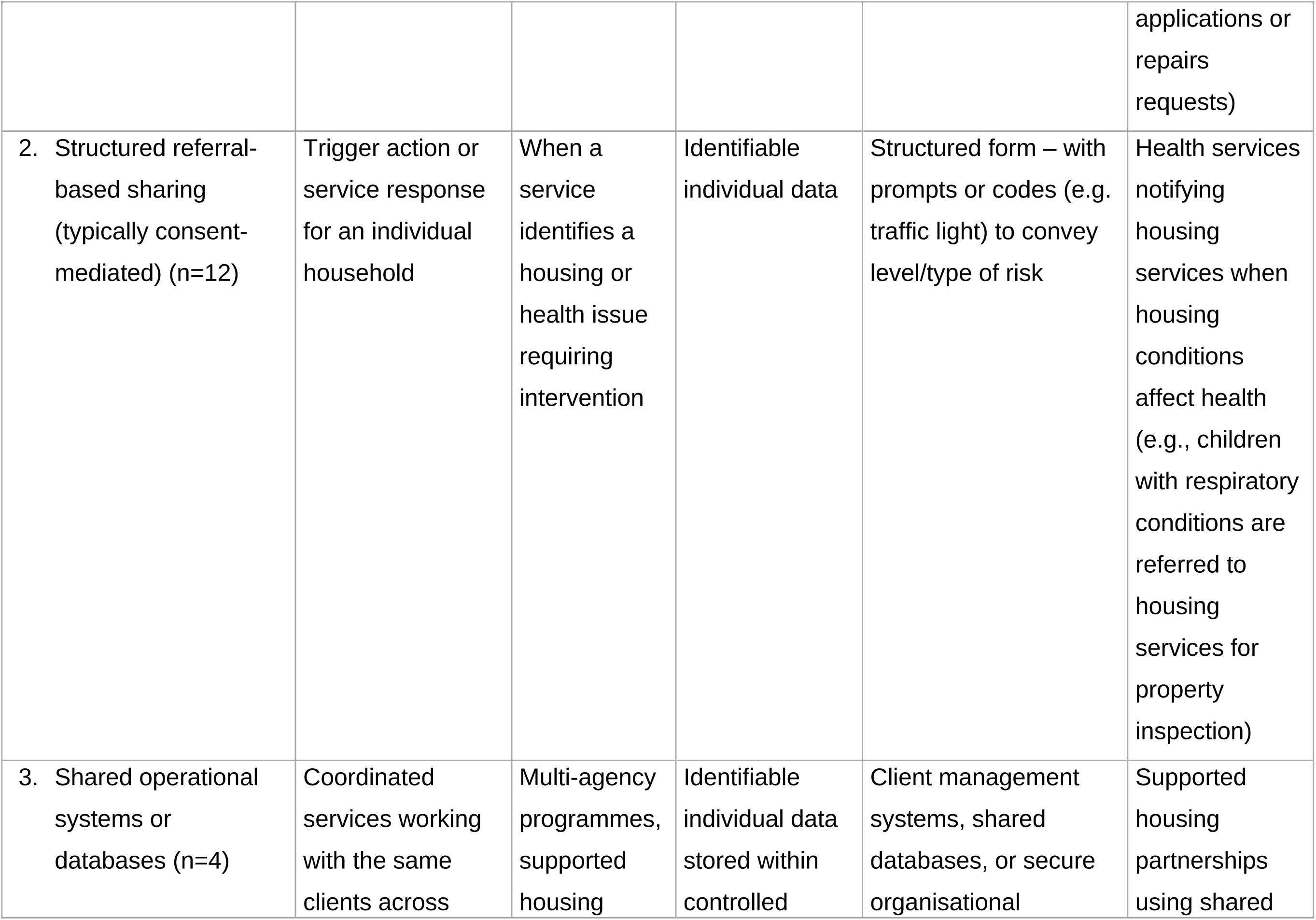
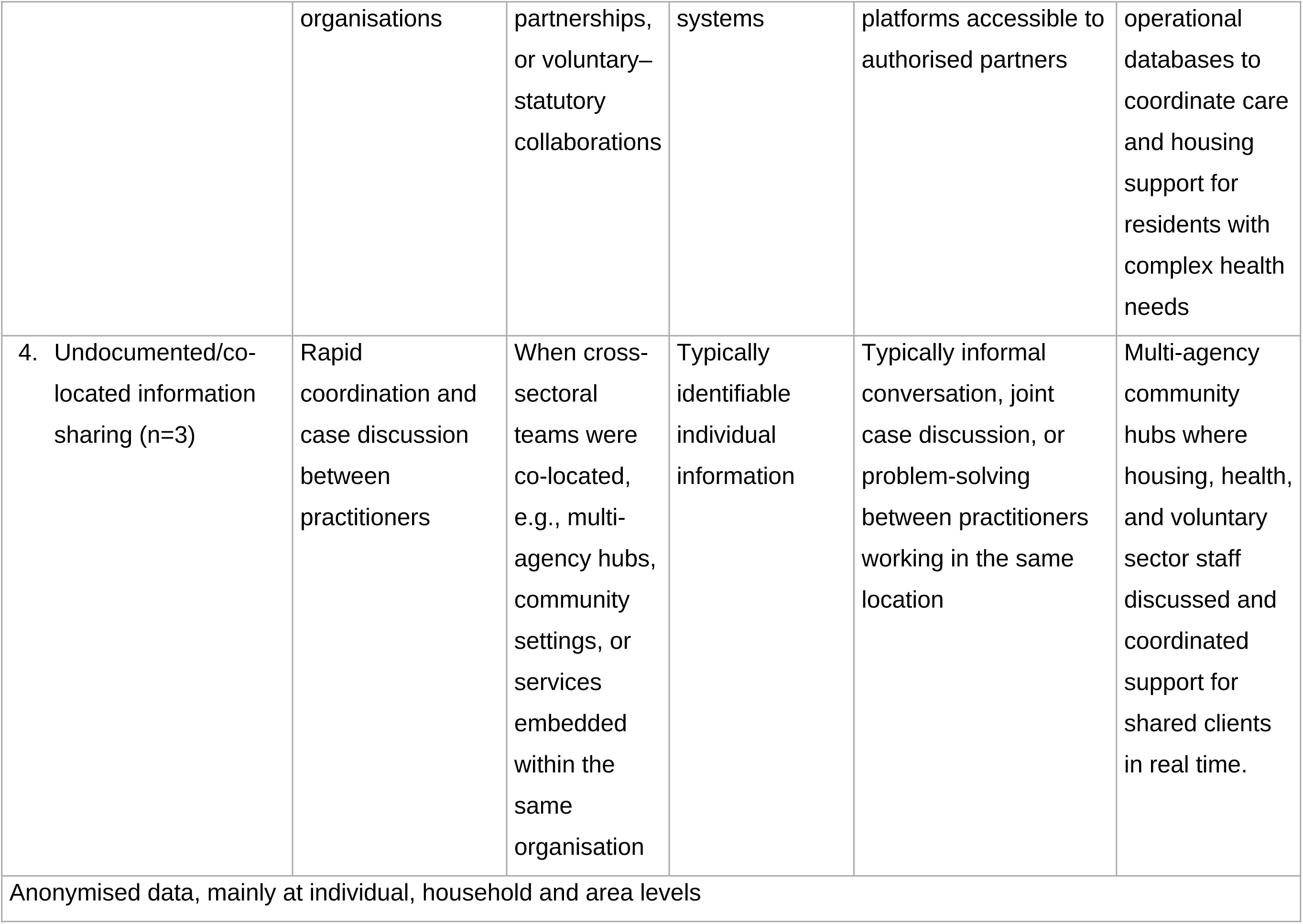
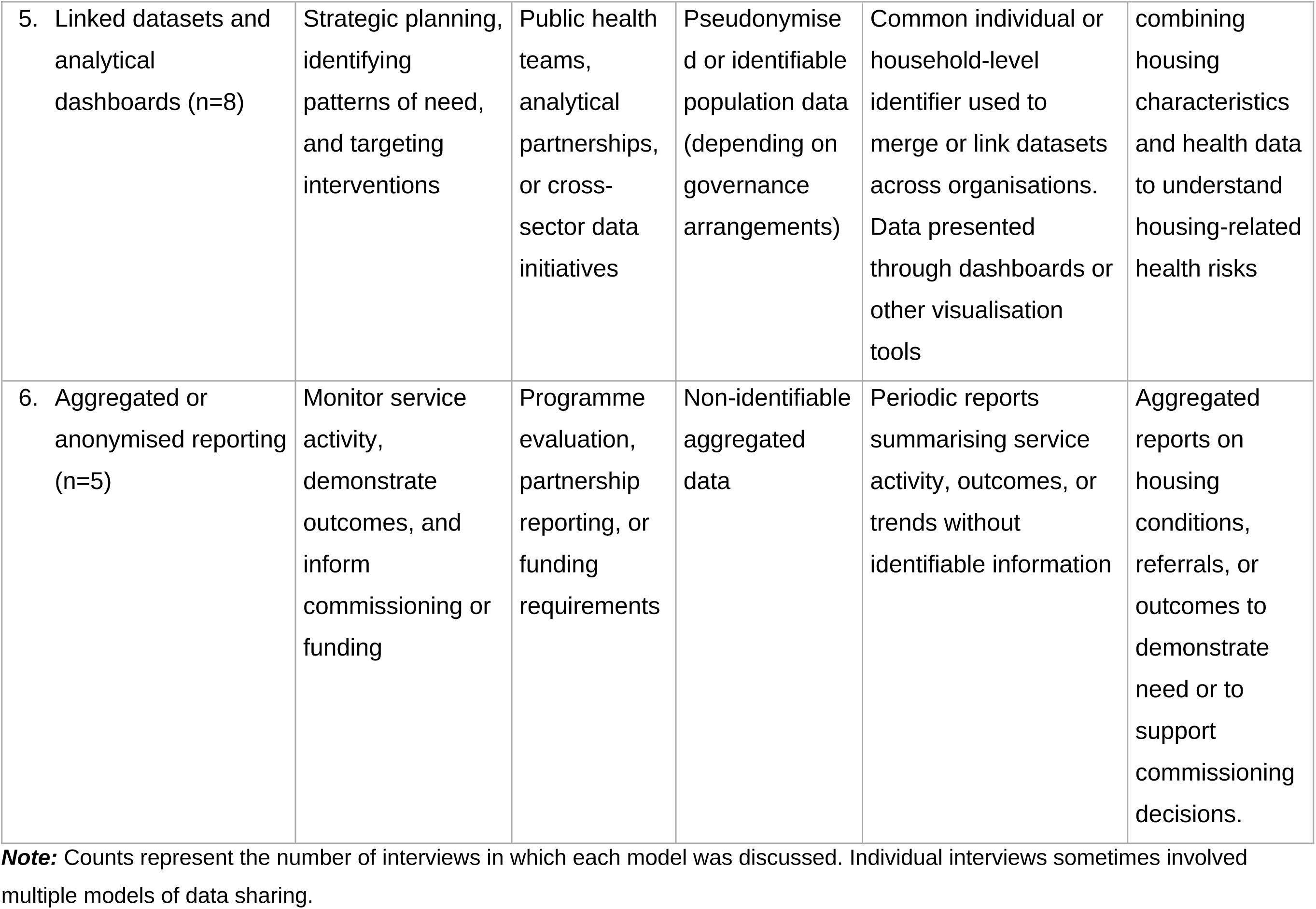
Models of cross-sector data sharing between housing and health organisations.

Analysis was conducted collaboratively within the research team. The researchers’ professional backgrounds and positions - as a researcher, a GP, member of a VCSE and a public health professional - influenced their interpretation of the data. For example, the public health researchers who have used patient and housing data for research purposes, had favourable perspectives on the benefits of using shared data appropriately, whereas the GP was conscious of the risks and professional responsibilities of clinicians in maintaining patients’ trust. Regular team discussions were used to refine coding, challenge interpretations and ensure that complexity and divergence within the data were retained. Attention was paid to identifying any contrasting perspectives across participants, tensions within individual accounts (e.g. simultaneous support for and concern about data sharing), and differences across models of data sharing.

## Results

The data comprised 27 interviews with participants from local authority housing and public health teams, housing organisations, NHS organisations, and VCSE sector organisations (Table 1). Participants from housing and local authority settings constituted the largest group (n = 14), followed by NHS organisations (n = 10) and VCSE sector organisations (n = 3). Just over half held strategic or senior roles (n = 15).

Data sharing initiatives most commonly mentioned by participants focused on housing quality (particularly damp and mould) and respiratory conditions (Table 2). Most involved the sharing of identifiable data at some stage (n = 23), although in some instances identifiable data informed anonymised population-level analysis.

Participants described data sharing as a practical means of coordinating action across housing, health and VCSE sectors. To progress coordination involved decisions about what information was relevant, who could access it, and how it should be acted upon. The findings are presented under four themes: (1) What counts as “data”, and what does data sharing look like, (2) Hopes and fears of data sharing, (3) What makes data sharing happen (and what hinders it), and (4) Impacts of data sharing.

Across all themes, different models of data sharing (Table 3) shaped how integration was enacted. A total of six models were identified with four related to individual identifiable data and two to anonymised data, typically at aggregate levels. The models were developed as analytical categories to capture dominant patterns of data sharing described by participants, but initiatives often combined multiple approaches or shifted between them over time. Enabling factors, barriers and impacts across models of data sharing were also identified (see Table 4).

**Table 4.** Distinct enabling factors, barriers and impacts across models of data sharing.

| Model of data sharing | Main enablers or facilitators | Main barriers or challenges | Typical impacts |
| --- | --- | --- | --- |
| 1. Unstructured referral-based sharing (VCSE-led, consent-mediated) | Trusting relationships with clients; flexibility; direct client involvement; contextual knowledge | Limited formal standardisation; reliance on individual advocacy; lack of system integration | Tailored support; stronger advocacy; increased transparency for clients |
| 2. Structured referral-based sharing | Clear referral pathways; standardised forms; alignment with workflows; defined responsibilities | Concerns about workload and referral volume; organisational capacity to respond; governance delays | Faster coordination; targeted inspections/interventions; improved cross-sector communication |
| 3. Shared operational systems/databases | Shared access to systems; ongoing collaboration; role integration | Governance complexity; uncertainty around permissions; dependence on specific staff roles | More coordinated case management; integrated frontline working |
| 4. Undocumented/co-located information sharing | Informal relationships; physical proximity; rapid communication | Fragility when staff leave; lack of accountability/documentation; unclear governance | Faster problem-solving; responsive coordination |
| 5. Linked datasets | Strong analytical | Complex governance processes; | Population health |
| and dashboards | capacity; organisational commitment;<br>governance agreements;<br>common identifiers | technical challenges; differing interpretations of legitimacy | management; targeting interventions; identifying inequalities; informing commissioning |
| 6. Aggregated or anonymised reporting | Lower governance burden; easier organisational sharing | Limited actionability at individual level; inability to coordinate direct intervention | Monitoring trends; demonstrating outcomes; informing funding/commissioning |

### Theme 1. What counts as “data”, and what does data sharing look like?

What counted as “data” varied across models of data sharing. Participants described a wide spectrum of materials and processes as data, ranging from highly structured digital datasets to informal conversations. Health-related data typically comprised NHS numbers, diagnostic codes and risk flags. Housing data included home inspection reports, photographs of damp and mould, environmental health assessments and address-level property classifications.

In unstructured referral-based data sharing (Model 1), described by VCSE organisations, data largely included lived experience narratives and contextual explanations of how housing affected clients’ wellbeing.

> “I think the key is really telling the story of how it’s impacting on the resident. So, it’s not this person has got this condition or that condition, it’s that this is how it’s impacting on their wellbeing, on their day-to-day life and also kind of the potential I guess risks to the resident if it’s not responded to. And sometimes that works. Most of the time it feels- And I guess the worrying thing is that when I have intervened, I’ll probably be more successful than not. But quite clearly, I can’t intervene with everybody. And it’s a worry that a resident, a patient cannot facilitate these sorts of outcomes directly themselves” (P017, VCSE)

Information was shared through free-text emails, often with clients copied into communication, allowing them to contribute documentation and retain oversight. Here, data were dialogical and co-produced rather than standardised. In contrast, structured referral-based models (Model 2) relied on predefined data fields, such as diagnostic risk categories or risk flags. These enabled clearer interpretation and action across organisations. For example, in a child asthma pathway, clinicians shared structured clinical information (e.g., diagnosis and severity) with housing teams to trigger property inspections and interventions.

> “For severe cases, it all goes through the same process and that makes it really nice and simple for clinicians and the housing team as well.” (P009, NHS/Health)

Shared operational systems (Model 3) involved access to patient/resident records across sectors, enabling coordination around individuals through controlled systems. Participants also described instances of sharing individual-level data even when there were no formal mechanisms (e.g. underpinning data sharing agreements, honorary contracts or processes) for recording what information existed (Model 4). Instances might include viewing a colleague’s computer screen when co-located or a telephone conversation for the same purposes as the more formalised case-based referrals.

At a population scale, linked datasets and dashboards (Model 5) combined housing and health data using common identifiers to generate population-level insights. Here, data were coded, linked and analytically derived, generating patterns and enabling targeted interventions. Finally, aggregated reporting (Model 6) involved summarised, identifiable and non-identifiable data used for analysis, monitoring and commissioning.

Participants also described the need to justify and define new ways of using existing data. In some cases, organisations developed formal “use cases” - structured written arguments submitted to governance bodies to justify sharing data in a particular way, typically as part of a Data Protection Impact Assessment process - to support approval for sharing data differently across sectors. Importantly, existing data sharing agreements could provide a foundation for subsequent use cases, with prior arrangements lending legitimacy and precedent to new requests and reducing the time required for approval.

> “The ICB [Integrated Care Board] had a data sharing agreement with our community’s team to do some winter pressures, winter funding, to do a winter project. And then once we signed up to the data toolkit, we did a use case on why we need to share this data in a different way.” (P013, Housing/LA)

This highlighted that data sharing was not simply a technical process of transferring information, but involved ongoing interpretive, organisational and governance work to establish what forms of sharing were considered appropriate and legitimate - work that could be eased where prior arrangements had already established trust and precedent.

Across models, data were actively constructed through decisions about what to include (or not include), how it would be structured, and who could access it. These decisions shaped how organisations could act on shared information and, therefore, how integration became possible.

## Theme 2. Hopes and fears of data sharing

Participants’ expectations of data sharing varied by model and were also characterised by tension, where even strong supporters expressed simultaneous optimism and concern. Their hopes and fears for data sharing reflected broader beliefs about the relationship between housing and health, as well as debates about the legitimacy of sharing sensitive information across organisational boundaries. While many saw data sharing as necessary to address interconnected housing and health problems, others emphasised the ethical and practical risks involved.

### 2.1. Hopes for improved coordination and action

Across models, participants viewed data sharing as essential for prevention, early intervention and reducing inequalities. In referral-based models, expectations centred on enabling timely and coordinated action. Structured pathways were seen as a mechanism for triggering interventions and reducing delays. For example, in an asthma pathway, housing was understood as a modifiable determinant of health, particularly in relation to damp, mould and fuel poverty.

> “They’ve always seen housing as an issue when they have children come in with problems… it was just an area that they could see you can make a difference with it if housing get involved.” (P009, NHS/Health)

In linked data models (Model 5), expectations focused on strategic capability, with combining datasets seen as enabling more proactive targeting and prioritisation. This included linking housing and health-related data to identify risk, and then to target households proactively, supporting interventions aimed at both housing conditions and wider wellbeing:

> “We linked the LIFT [low income family tracker] data set with the [local housing] dataset to identify… households… at high risk… and… target these households… to have that conversation with them”. (P027, Housing/LA)

Across settings, participants framed the benefits of data sharing for residents, in reducing duplication and improving residents’ experience of support. It was noted that residents often had to repeatedly explain their circumstances to different services, which could be exhausting and frustrating:

> “I think it’s more that residents actually…have to go through things again, and they do report feeling tired of that.” (P007, NHS/Health)

### 2.2. Fears about misuse, intrusion and unmet need

Concerns varied depending on how data were shared. In referral-based models, a key fear was the obligation to act on identified need without sufficient capacity. Sharing information about poor housing conditions or health vulnerabilities might reveal a need for rehousing or major repairs when suitable housing or resources were not available. There were concerns that identifying need without sufficient resources to respond could expose unmet need or create expectations that could not be fulfilled. However, these concerns were not always realised in practice:

> “I think that was the fear… that they were going to be bombarded with loads of referrals and that’s not been the case.” (P009, NHS/Health)

Participants also raised concerns about trust and legitimacy. Data sharing could be perceived as intrusive if residents believed information was accessed without their knowledge or involvement:

> “If somebody came to your house… and there was any indication that they already knew the health condition of the people in the house, how would you feel if that data had been shared with a party that didn’t need to know?” (P014, NHS/Health)

VCSE sector participants emphasised the importance of limiting what information was collected or shared, as well as who had access to it, to reduce the risk of harm to residents. For example, one participant reported deliberately avoiding collecting or sharing ethnicity information unless it was clearly necessary because of fear of discrimination.

Maintaining public trust was seen as fundamental:

> “Part of the reason common law duty exists is the confidence that the public have in services. So, when you tell your GP something… In order for that service to be effective it has to have the confidence of those who it cares for…if that confidence is lost it does break down the possibility of people sharing basically their health data with a health professional…That’s a risk is that we lose confidence in public services if we don’t do this correctly” (P014, NHS/Health)

These concerns illustrate the “dark logics” of data sharing: the recognition that information sharing can both enable support and create new risks if used inappropriately:

> “… we all believe that it can be abused and to discriminate. It can be used for the good, but it can also be used for abuse as well.” (P016, VCSE)

Across models, participants differed in how they interpreted what was legitimate or proportionate. For some participants, strict interpretation of governance rules was necessary to protect residents’ confidentiality and maintain trust. Others suggested that organisations could interpret these rules overly cautiously.

These differences were not only between organisations but sometimes within the same individual. Several participants considered how they or their own family members might respond if their data were shared in the ways they were describing, grounding their professional judgements in personal as well as organisational terms:

> “…in an ideal world, it would be great if a lot of information would come together…it would make a very effective system, yes. But who would you authorise to have access to such sensitive information from all aspects, you know? Because that would mean that you expose a person’s everything, you know, to someone. And I don’t know how comfortable I would be, you know, if somebody would, in a more or less uncontrolled way, have an insight into my data, you know, and stuff like that, and like how many abuses can happen because of that” (P022, Housing/LA)

Thus, expectations about data sharing were shaped not only by perceived benefits and risks, but by judgements about whether organisations had the capacity to act on the information shared and ongoing negotiation of legitimacy.

## Theme 3. What makes data sharing happen (and what hinders it)?

In this section, we focus on enablers, given the evidence gap in this area, but highlight distinct barriers identified across models (Table 4).

### 3.1. Enablers that supported implementation

Several factors allowed data sharing initiatives to be implemented and sustained. In referral-based models, working with partners to agree structured forms, including the use of standardised fields and risk categorization (e.g., traffic light systems) reduced ambiguity and supported implementation:

> “So, we started off by very clearly-By working with each partner to say, right, I will give you name, address, postcode and a little bit of information about that cohort as well…I could have given them so much more, but it is an absolute no. They just need to know the basics. We agreed a common purpose to say this data is for you to go and talk to these households. You don’t necessarily need to know everything about the household.” (P027, Housing/LA)

Referral-based models were also more readily embedded where they aligned with existing clinical workflows (e.g. a clear referral route for clinicians and an understanding of the role of housing teams), which reduced uncertainty and duplication. Participants also emphasised that successful implementation depended on understanding how partner organisations worked in practice. Participants described how greater understanding of each other’s systems and responsibilities helped develop referral pathways that were both acceptable and feasible across organisations.

As one participant explained:

> “Doctors, nurses, medical people like pathways… We’re process-driven animals… We didn’t want to bring anything new in. We wanted to give them something on a platform that they were very familiar with and that they understood as well.” (P018, NHS/Health)

Similarly, one participant who had previously worked as a nurse reflected on how limited understanding of housing could hinder collaboration:

> “…a nurse who doesn’t know how complicated housing is, because I didn’t know until I came onto the council and flipping heck… it’s a minefield.” (P018, Housing/LA)

In linked data models, progress depended on sustained negotiation, governance alignment and organisational commitment. For example, temporary changes to data sharing permissions led to a regional linked dataset integrating NHS and local authority data, which led to the development of a dashboard allowing authorised health professionals to identify high-need households and generate referrals to housing partners. Importantly, housing associations did not directly access identifiable health data; instead, health teams mediated referrals:

> “We ingested the housing data into the xxx database and… create[d] a dashboard where we could identify… who can legitimately see the health data.” (P014, NHS/Health)

Co-location and informal interaction acted as important enablers of data sharing across all models. Participants described how being physically or organisationally embedded within the same environment facilitated real-time communication, joint problem-solving and the sharing of contextual information that might not be captured through formal systems. In these settings, data sharing sometimes occurred through conversation, case discussion or direct access to shared systems, rather than through formalised processes (Model 4).

> “…I mean it’s hard work as it is because building relationships between housing and health is tricky at the best of times because both are really, really busy with conflicting priorities…I think it needs to be part of who you are” (P005, Housing/LA)

However, these forms of informal sharing were dependent on personal relationships and could be difficult to sustain. Collaboration at times relied on specific individuals, and when those individuals left, connections were weakened or lost:

> “It’s that relationship with one person and when that person leaves… you kind of lose those links.” (P015, Housing/LA)

Beyond personal relationships, these forms of informal sharing were also dependent on organisational structures (e.g., joint appointments, housing and health networks or board) that were not always sustainable and were vulnerable restructuring or funding cuts.

Importantly, barriers and enablers did not operate independently. Governance processes initially slowed implementation but later enabled legitimacy. Similarly, concerns about workload were mitigated where structured processes clarified roles and limited demand. As one participant reflected,

> “ It can’t go through unless it’s got all that detail in it. I mean it’s not serving, the design of it has to include all the different parties and I know it takes time, and I just keep saying we do this work at the outset. If we do it at the beginning, it will pay off later, I promise you, I just know it will. But if you don’t address it now, you’ll lose it and then you’re losing pieces that could be really helpful.” (P004, Housing/LA)

### 3.2. Barriers to data sharing across models

In referral-based models, particularly those involving clinicians, challenges related primarily to additional workload:

> “Each of those things have just taken months and months” (P014, NHS/Health)

In contrast, linked data models were more likely to be constrained by governance complexity and differing interpretations of data protection.

> “It can mean things get shared that maybe shouldn’t [also an example of a fear being realised] …or it can mean things don’t get shared because everyone’s a little bit risk averse when actually they could be shared and they should be shared” (P020, VCSE/Housing)

Formal data-sharing agreements were often time-consuming to establish and not always accessible to smaller housing providers and VCSE organisations, who often lacked specialist data protection support:

> “So we’ve got information sharing agreements in place with all of those different authorities, which was in some cases really almost surprisingly easy. In others it was almost surprisingly and painfully slow and difficult.” (P019, NHS/Health)

## Theme 4. Contribution of data sharing to changes for residents and/or services

### 4.1. Differences in impact by model

The types of impacts of data sharing described by participants also differed across models of data sharing (Table 4). Impacts were shaped by: the availability of evidence; the time required for implementation; and wider structural constraints (including funding and governance arrangements). Where shared operational systems or linked datasets were used, data sharing supported more strategic forms of coordination. In these settings, housing providers were sometimes repositioned as contributors to health-related activity. The rapid development of linked datasets during the pandemic appeared to strengthen the perceived value of cross-sector data sharing, with one participant noting that “the pandemic accelerated the thinking and the possibility”.

Changes in frontline practice and individual-level outcomes from referral-based models included targeted environmental inspections, fuel poverty referrals and minor home adaptations. As one participant noted, referral pathways provided “more of a reason to go and visit that property and check the condition of it…” (P009, NHS/Health), offering a clearer basis for action. In some cases, having access to health-related information enabled housing providers to prioritise interventions based on residents’ specific vulnerabilities, supporting more proactive and personalised responses.

In VCSE sector settings, unstructured referral-based sharing supported more tailored and responsive forms of support, particularly for individuals with complex or overlapping needs. In this model, VCSE advocates could help clients to see, contribute to and sometimes shape what information was shared about them, which was seen as enhancing transparency and engagement.

### 4.2. Broader system effects and limitations

Participants described shifts in how professionals understood their roles and the relationship between housing and health. Data sharing led to opportunities for collaboration and developing new forms of joint working (through shared operational systems):

> “It’s turned into a much more collaborative approach… I can now go to [VCSE sector organisation] and ask what they can tell me about an area… the connections are there” (P027, Housing/LA)

In some cases, data sharing broadened perspectives, enabling healthcare staff to recognise housing as a determinant of health and to consider how their work could contribute to addressing inequalities. Access to linked data also enabled some services to identify and respond to inequalities more explicitly, for example by targeting interventions toward households experiencing both poor housing conditions and health vulnerability.

> “I am now working with departments and external organisations about how best we can use this [housing] data… see what else it can do, particularly how we can conjoin it with other data sets within the council to just add a bit more information about what we know of a household and to shape some campaigns.” (P027, Housing/LA)

Immediate positive outcomes were reported for some residents, including improved housing conditions, more coordinated support and, in some cases, reductions in hospital admissions:

> “In terms of the number of children that have come through this pathway, over a 12-month period there was only one red case… the family got rehoused in that situation. It reduced the number of admissions to hospital as well…” (P009, NHS/health)

However, impacts were not always clear and sustained. In some cases, it was too early to determine outcomes. In others, evidence of benefit from pilot projects did not translate into wider programmes:

> “There is a wealth of evidence out there based off pilot projects where obviously a group of homes have been improved, and the evaluation has been done. And therefore, the evidence of improved health of the residents has been clearly identified but then it doesn’t go any further…the wider improvement programmes can’t go ahead because there’s no money to do them.” (P015, Housing/LA)

## Discussion

This interview study explored how data sharing between housing and health organisations operates in practice across multiple contexts in the UK. Data sharing initiatives rarely followed a linear trajectory from pilot to scale. Instead, they required active meaning-making and legitimation, evolving through adaptation, recalibration and, in some cases, intentional limitation. Integration was produced through agreeing together what counted as relevant data and negotiating legitimacy in the contexts of routine activity in each organisation.

### Methodological considerations

This study provides one of the first empirical accounts focused specifically on cross-sector data sharing between housing and health, addressing a gap in the literature [10, 16]. It draws on perspectives across multiple sectors and roles, offering insight into both system-level and frontline processes.

The study has several limitations. The sample was weighted toward housing and local authority participants, particularly those in senior or strategic roles. While this enabled insight into the initiation and governance of data sharing, it may underrepresent frontline perspectives and the practical challenges of implementation. Furthermore, the cross-sectional nature of the study captured accounts of data sharing initiatives at one time rather than tracing their development over time. As a result, findings lack insight into longer-term sustainability or outcomes. Invitations to take part in the study were shared widely, seeking individuals with experience of making progress with data sharing across sectors. While most individuals that expressed an interest in taking part agreed to be interviewed and recorded, we recognise that negative experiences of, and the barriers to, data sharing are not captured as fully as enablers. The categorisation of data sharing initiatives into distinct models represented an interpretive analytic process based on our broad framing of “data sharing”. Alternative interpretations may have classified examples differently.

We also reflect on the applicability of Normalisation Process Theory as a sensitising and organising framework. It helped to foreground the work required to make data sharing meaningful (coherence), legitimate (cognitive participation), operational (collective action) and appraised over time (reflexive monitoring). However, the findings also suggest that data sharing involves broader ethical and relational dimensions that extend beyond implementation processes alone. NPT was less able to capture the ethical tensions and wider system constraints that shaped decisions about whether and how data should be shared.

### Comparisons with other literature

Policy, legislative and technological developments in data sharing within healthcare have focused on increasing data standardization as a key enabler to data integration. For example, in England, the Health and Social Care Act (2012), followed by the Health and Care Act (2022) and the Data Use and Access Act (2025) have increased requirements for standardization of health data by public services and now IT suppliers [17]. Tweed et al’s review and recommendations for data sharing across Scotland also identifies the need for data standardisation across both health and non-health datasets [10]. While data standardisation is necessary, the focus on it may imply that data sharing is a technical exercise, and that the goal of specific initiatives is to identify a single, optimal model that can be scaled up. Our identification of distinct models of data sharing - each associated with different forms of integration - challenge the applicability of these assumptions to cross-sector data sharing in a service context. In our study, integration through data sharing operated across multiple scales, from individual-level unstructured and structured mechanisms such as referral pathways to population-level datasets. Integration depends on the combination of different models, each enabling different types of action. Importantly, the boundaries between these models were not always fixed or mutually exclusive. In practice, initiatives often combined multiple forms of data sharing or shifted from identifiable, individual data to aggregated, analytical approaches over time.

The study also highlights that data sharing is actively negotiated. Participants did not perceive data sharing as inherently beneficial; rather, they weighed potential gains against risks, including concerns about privacy, misuse and loss of trust. Importantly, these tensions were not only between organisations but often within individuals, who expressed both optimism and concern. This reflects the complexity of data sharing as both a technical and ethical practice.

Existing literature has identified governance and organisational barriers to data sharing [1,8]. Across sectors, others have found that data sharing complexities are compounded by working across multiple different organisations, each with different competing demands and priorities; and sequential and interdependent governance processes which exacerbated delays and led to unpredictable timelines [10]. Our findings suggest that such factors are not fixed constraints. Instead, they are interpreted and negotiated in context, shaping how data sharing is enacted. Decisions to limit or adapt data sharing were not always insurmountable barriers, but constituted active forms of ethical and professional judgement, through which people balanced competing obligations to act, protect and avoid harm. Rather than obstacles to overcome, governance restraints may instead play a role in shaping what forms of integration are possible and appropriate and how to ensure it is done safely and ethically.

### Conditions for implementation

In common with the wider literature [1,10], we also found that relational factors were critical to achieving data sharing. As Sheppard et al [18] reported, we found that pre-existing partnerships, named key contacts, informal interaction and co-location supported coordination by enabling real-time communication and shared problem-solving and faster negotiation of arrangements. Our findings extend the general principle of the importance of relational factors by illustrating how organisational relationships were important to specific data sharing practices beyond merely the formation of personal connections. We identified a range of inter-organisational partnership arrangements such as joint appointments, funded housing and health networks or boards that facilitated formation and maintenance of data sharing through ensuring the right individual were connected, being able to translate terminology across sectors and supporting transfer of knowledge about often implicit routines, workflows and constraints across sectors. In response, we found that structured referral pathways were more readily embedded where they fitted within routine practice and clarified roles and responsibilities. However, these could be fragile when organisational restructures and funding cuts led to cancellation of these arrangements.

Further, whilst VCSE sector organisations were central to many initiatives, they were sometimes not included in formal data-sharing arrangements, reflecting wider challenges within integrated care systems, where non-statutory actors may be essential to delivery but are not always recognised within governance structures.

### Implications for policy and practice

Current UK policy and legislative changes position data sharing as central to integration, with a strong emphasis on standardised digital infrastructure and interoperability [17]. This risks underestimating the relational, organisational and ethical work required to make data sharing effective in practice. Further, as different models of data sharing support different forms of integration, individuals seeking to develop data sharing arrangements could consider which models are applicable in their context and whether to combine approaches that enable both individual-level coordination and population-level planning.

## Conclusion

Data sharing between housing and health is often framed as a technical challenge. This study suggests a more nuanced understanding. Integration is not simply enabled by data sharing; it is produced through the practices of defining, negotiating and embedding data sharing across organisational boundaries.

Supporting integration therefore requires attention not only to systems and data, but to the work required to make them meaningful, legitimate and actionable in practice.

## Supplementary material

Supplementary File 1: Interview topic guide

Supplementary File 2: Rapid appraisal template

Supplementary File 3: Initial NPT codebook with examples of coded text

Supplementary File 4: COREQ (Consolidated Criteria for Reporting Qualitative Research) checklist

## Ethics statement

Ethical approval was granted by the UCL Life and Medical Sciences Research Ethics Committee (reference 0128). All participants provided informed written consent prior to taking part. All relevant ethical guidelines were followed.

## Data availability statement

The data that support the findings of this study consist of interview transcripts containing potentially identifiable information about participants. In line with the consent provided by participants, these data are not publicly available. Requests to access the data may be considered by the corresponding author on reasonable request.

## Acknowledgements

We would like to thank the following for their advice on the study and in brokering contact with potential research participants: Liz Parsons, Director of Community Investment, Amplius Housing Association; Sarah Rodgers, Professor of Health Informatics, University of Liverpool; Laura Saksena, Chief Executive of HealthWatch Islington. We also express sincere thanks to Kerry Littleford, Public Health Specialty Registrar, Royal Free NHS Foundation Trust, for her detailed review of an earlier draft of this manuscript.

