## Supplementary file 1 for "Shared Data As a Mechanism for Integration between Housing and Health Sectors: A Qualitative Interview Study in the UK"

**Rapid appraisal sheet:**

**Interview ID:**

**Data flow**

**Brief description**

**Direction**

- Housing → health
- Health → Housing
- Health ↔ Housing

**Identifiable data?** Yes/no

**Approval for sharing**

- Patient/resident consent sought on case by case basis?
- DPIA or other local approval process for sharing across a population?
- National eg CAG/Section 251?
- Other

**Data sharing outputs**

- Identifiable
- Individual but anonymised
- Aggregated

- Dashboard
- Referral form
- Other

**Condition**

- No specified
- Respiratory
- CVD
- Mental health
- Other
- Not specified
-

**Housing problem**

- Overcrowding

- Damp and mould
- Housing condition, e.g. repairs
- Housing demand – homelessness, temporary accommodation
- Other
- Not specified

**Population group**

- Children
- Older adults
- Homelessness
- Other
- Not specified
