## Supplementary file 2 for "Shared Data As a Mechanism for Integration between Housing and Health Sectors: A Qualitative Interview Study in the UK"

Supplementary File 1: COREQ checklist

### Consolidated criteria for reporting qualitative studies (COREQ): 32-item checklist

Developed from:

Tong A, Sainsbury P, Craig J. Consolidated criteria for reporting qualitative research (COREQ): a 32-item checklist for interviews and focus groups. International Journal for Quality in Health Care. 2007. Volume 19, Number 6: pp. 349 – 357

| Item No | Guide Questions/Description | Reported on Page # |
|---|---|---|
| Domain 1: Research team and reflexivity | | |
| Personal Characteristics | | |
| 1. Interviewer/ facilitator | Which author/s conducted the interview or focus group? | Methods, page 6 “Data collection” |
| 2. Credentials | What were the researcher’s credentials? E.g., PhD, MD | Title page |
| 3. Occupation | What was their occupation at the time of the study? | (Noted in analysis section) Page 8 |
| 4. Gender | Was the researcher male or female? | Not added |
| 5. Experience and training | What experience or training did the researcher have? | Page 6 |
| Relationship with participants | | |
| 6. Relationship established | Was a relationship established prior to study commencement? | Page 6 |
| 7. Participant knowledge of the interviewer | What did the participants know about the researcher? e.g. personal goals, reasons for doing the research? | Information sheet shared ref protocol page 6 |
| 8. Interviewer characteristics | What characteristics were reported about the interviewer/facilitator? e.g. Bias, assumptions, reasons and interests in the research topic | (analysis section when discussing the professions of the research team) Page 9 |

| Item No | Guide Questions/Description | Reported on Page # |
|---|---|---|
| Domain 2: study design | | |
| Theoretical framework | | |
| 9. Methodological orientation and Theory | What methodological orientation was stated to underpin the study? e.g. grounded theory, discourse analysis, ethnography, phenomenology, content analysis | (Normalisation process theory) Page 3 |
| Participant selection | | |
| 10. Sampling | How were participants selected? e.g., purposive, convenience, consecutive, snowball | (Purposive and then snowball) Page 6 |
| 11. Method of approach | How were participants approached? e.g., face-to-face, telephone, mail, email | Page 6 |
| 12. Sample size | How many participants were in the study? | 27 Page 9 |
| 13. Non-participation Setting | How many people refused to participate or dropped out? Reasons? | Data collection Page 7 |
| 14. Setting of data collection | Where was the data collected? e.g., home, clinic, workplace | (Online via MS Teams) Page 7 |
| 15. Presence of nonparticipants | Was anyone else present besides the participants and researchers? | Page 7 |
| 16. Description of sample | What are the important characteristics of the sample? e.g. demographic data, date | (Level of profession, area of work) Page 10 |
| Data collection | | |
| 17. Interview guide | Were questions, prompts, and guides provided by the authors? Was it pilot tested? | (Topic guide linked) Page 7 |
| 18. Repeat interviews | Were repeat interviews carried out? If yes, how many? | (No) |
| 19. Audio/visual recording | Did the research use audio or visual recording to collect the data? | (Audio) Page 7 |
| 20. Field notes | Were field notes made during and/or after the interview or focus group? | Page 7 |

| Item No | Guide Questions/Description | Reported on Page # |
|---|---|---|
| 21. Duration | What was the duration of the interviews or focus group? | (45-60 mins)<br>Page 7 |
| 22. Data saturation | Was data saturation discussed? | Sought information power and diversity of sample rather than saturation (see page 6) |
| 23. Transcripts returned | Were transcripts returned to participants for comment and/or correction? | (No) |
| **Domain 3: analysis and findings** | | |
| **Data analysis** | | |
| 24. Number of data coders | How many data coders coded the data? | Page 8 |
| 25. Description of the coding tree | Did the authors provide a description of the coding tree? | Reference to coding framework in supplementary data page 5 |
| 26. Derivation of themes | Were themes identified in advance or derived from the data? | Themes informed in advance by use of mid-range theory (NPT) then informed by the data.<br>Page 8 |
| 27. Software | What software, if applicable, was used to manage the data? | NVivo<br>Page 8 |
| 28. Participant checking | Did participants provide feedback on the findings? | (No) |
| **Reporting** | | |
| 29. Quotations presented | Were participant quotations presented to illustrate the themes/findings? Was each quotation identified? e.g., participant number | Quotations by ppt number (Results, pp19-30) |

| Item No | Guide Questions/Description | Reported on Page # |
|---|---|---|
| 30. Data and findings consistent | Was there consistency between the data presented and the findings? | Yes |
| 31. Clarity of major themes | Were major themes clearly presented in the findings? | Yes |
| 32. Clarity of minor themes | Is there a description of diverse cases or a discussion of minor themes? | Page 8 |
