## Supplementary file 3 for "Shared Data As a Mechanism for Integration between Housing and Health Sectors: A Qualitative Interview Study in the UK"

This is an adaption of May et al.'s (2022) normalisation process theory coding manual, which set out NPT constructs using a Context-Mechanism-Outcome framework, defined them and provided examples.

<table>
<tr><th>NPT construct</th><th>Sub-construct</th><th>Description</th><th>Examples please add an annotation for key quotes why it fits the code/is important to answer the research question)</th></tr>
<tr><td rowspan="4">Implementation contexts: Contexts are patterns of social relations and structures that unfold over time and across settings. They make up the implementation environment.<br><br>There are many CFIR outer/inner setting (https://cfirguide.org/constructs/) dimensions.<br><br>- Local attitudes (sociocultural values around shared responsibility)<br>- Local conditions (technological, political, economic<br>- Partnership and connections<br>- Policies and laws<br>- Financing, e.g. funding available to implement and deliver the intervention</td><td>Strategic intentions</td><td>Description: How do contexts shape the formulation and planning of interventions and their components?</td><td rowspan="4">External:<br><br>Internal/System:</td></tr>
<tr><td>Adaptive execution</td><td>Description: How do contexts affect the ways in which users can find and enact workarounds that make an intervention and its components a workable proposition in practice?</td></tr>
<tr><td>Negotiating capacity</td><td>Description: How do contexts affect the extent that an intervention and its components can fit, or be integrated, into existing ways of working by their users?</td></tr>
<tr><td>Reframing organisational logics</td><td>Description: How do existing social structural and social cognitive resources shape the implementation environment?</td></tr>
<tr><td>Coherence: How do people work together to understand and plan</td><td>Differentiation</td><td>Description: How did people (leaders and on the</td><td></td></tr>
</table>

| | | |
|---|---|---|
| the activities that need to be accomplished to put an intervention and its components into practice? | | ground staff) perceive that things would be different?<br>What did actors to do initiate, develop and progress plans for sharing data? |
| | **Communal specification** | **Description:** How did staff collectively view the rationale behind sharing data?<br>What did they think were the reasons others had for supporting/opposing it? |
| | **Individual specification** | **Description:** How did people individually understand what work sharing data would require of them? |
| | **Internalisation** | **Description:** How did people <u>construct</u> potential value of sharing data for <u>their work</u>? |
| **Cognitive participation:** How do people work together to create networks of participation and communities of practice around sharing data? | **Initiation** | **Description:** How did key individuals drive the implementation of sharing data?<br>How were stakeholders brought together in the planning, delivery or monitoring of sharing data? |
| | **Enrolment** | **Description:** How were people introduced to, and involved in, the implementation of sharing data? |

| | | |
|---|---|---|
| | **Legitimation** | **Description:** How do people agree that sharing data is the right thing to do and should be part of health/housing delivery? |
| | **Activation** | **Description:** How do people continue to support the delivery of sharing data? |
| **Collective action:** How do people work together to enact data sharing? | **Interactional workability** | **Description:** How do people do the work required by sharing data? |
| | **Relational integration** | **Description:** How does using sharing data affect the confidence that people have in each other? |
| | **Skill-set workability** | **Description:** How is the work of sharing data appropriately allocated to people? 34ex |
| | **Contextual integration** | **Description:** How is the work of sharing data supported by host organisations? |
| **Reflexive monitoring:** How do people work together to appraise data sharing? | **Systematisation** | **Description:** How do people access information about the effects of sharing data? |
| | **Communal appraisal** | **Description:** How do people collectively assess sharing data as worthwhile? |
| | **Individual appraisal** | **Description:** How do people individually assess |

| | | |
|---|---|---|
| | | sharing data as worthwhile? |
| | **Reconfiguration** | **Description:** How do people modify their work in response to their appraisal of sharing data? |
| **Implementation outcomes:** The practical effects of implementation mechanisms at work. | **Intervention performance** | **Description:** What practices have changed as the result of sharing data being operationalised, asenacted, reproduced, over time and across settings? |
| | **Relational restructuring** | **Description:** How has working with sharing data changed the ways people are organised and relate to each other? |
| | **Normative restructuring** | **Description:** How has working with sharing data changed the norms, rules and resources that govern action? |
| | **Sustainment (normalisation)** | **Description:** How has sharing data become incorporated in practice? |

Other key concepts to code

| | |
|---|---|
| Data flows created | |
| Disease or healthcare issue | Mental health<br>Respiratory health, asthma<br>Cardiovascular disease<br>Vaccination<br>Other<br>Not mentioned/vague |

| | |
|---|---|
| Housing issue | Homelessness<br>Temporary accommodation<br>Overcrowding<br>Damp and mould<br>Disrepair<br>Other<br>Not mentioned/vague |
| Population | Children<br>Older people<br>Homeless<br>Social housing tenants<br>other<br>Not mentioned/vague |
| Sectors between which data are shared | primary care<br>acute care<br>mental health<br>housing provider<br>VCS<br>Other |
| Potential barriers | Information governance<br>Patient concerns<br>Staff concerns<br>Understanding data<br>Technical barriers |
| Enablers | Individual<br>Organisational<br>Cultural<br>Technical |
