## Supplementary file 4 for "Shared Data As a Mechanism for Integration between Housing and Health Sectors: A Qualitative Interview Study in the UK"

Ethics number: 0128

**Title of Study:** Priorities, barriers and opportunities for using shared housing and health data to reduce health inequalities (nested study within the DASHH programme)

**Short title:** Priorities, barriers and opportunities for shared housing and health

### TOPIC GUIDE

V1.4 June 2025

- ***Confirm consent has been signed and agreement to record***
- *Check in on how long they have got. If time is short prioritise questions in yellow and consider dropping those in grey.* ***[NB prioritisation to be finalised through piloting]***
- *Press record*

Introduction: This project is part of a larger programme of work called DASHH (DAta Sharing for Housing and Health) that seeks to understand how sharing information responsibly between housing and health services could be used to benefit residents or patients and reduce health inequalities. **Information might include individual or population-level data on resident or patient records, insights gained from discussions with patients or residents.**

| **1. YOUR ROLE** |
| --- |
| Possible prompts<br>- Directorate/team?<br>- Involvement/responsibility for housing/health<br>- Experience of information sharing to date (successful or not?) |
| **2. EXPERIENCE OF INFORMATION SHARING TO DATE** |
| Could I ask about examples of information sharing between housing and health that take place in your service/organisation (across departments or organisations)?<br>At what stage is this?<br>What information has or will be shared and with whom? |
| **3. VIEWS ON SHARING INFORMATION (DATA) BETWEEN HOUSING AND HEALTH SERVICES** (COHERENCE/COLLECTIVE ACTION) |
| - When did you **first consider** the possibility of sharing information in this way? How /or when was it proposed to you?<br>- What did you see as the **opportunities/potential uses** of it in general?<br>- *Prompts; for residents/patients? For decision making? Service planning or provision? For workflows?* |

- What about the **risks? Limitations**? Potential harms
- *Prompts; Privacy/confidentiality? Workload?*

  How do you see it could **affect inequalities**?
  *Prompts: opportunity to measure/flag inequalities, benefits to people that can't or aren't ready to report problems themselves, a risk to people with uncertain migration status?*

- What about your colleagues? What did they think about it? Why did they think that? *Prompt for range of views amongst colleagues; explore if particular professional groups had particular reactions.*

- What, in your opinion do you think influenced **the [x organisation's] decision** to go ahead with it?

*Possible prompts: e.g.* How does it fit with the existing ethos/culture of your service?

4. **IMPACTS ON YOU, SERVICE, TEAM (**Collective action)

*If data sharing has started…*

Can we turn now to the impact that information sharing has on your work, and the work of colleagues.

How has it changed your **interactions with residents/patients / consultations AND/OR discussions with colleagues/senior management** ? How does that feel?

*Prompts: Effects on ease and effectiveness of consultations?*

In your opinion how do you think data sharing changes the **skills** you need to do your job? If so, how, and how does that feel?

- Did you, or your colleagues, need training before you could use the data? How hard / easy did you find it to learn to use the data? Did you receive any training?
- Conversely are there skills that you used to use before having access to these data that you don't need (or need as much) since then?

Has data sharing had any impact on the **division of work or the relationships between different professional groups** or services? If so, how, and why?

- *Possible prompt: Can you talk me through an example of something that you personally did/do you do differently?*

Is there anything special or different about **the service you work in** that made implementing data sharing particularly hard or easy? If so, what / why?

*Possible prompts:*

- *Data quality or local platforms*
- *Sharing agreements*
- *Personal relationships*
- *service priorities or recognised problems*
- ethics/culture around data

5. **REFLECTIONS AND CHANGES (**Reflexive monitoring)

| |
|---|
| *If data sharing has started…*<br>- What **changes have been made** since you started? Can you talk us through these changes and explain why they were made? Why? Would like to make further changes? Why is that? *Prompts:*<br>- *Type of data: population, granularity,*<br>- *Frequency of sharing/updating*<br>- *Access/who it's shared with and how*<br>- *How it's used*<br><br>- Some people have mentioned **changes to the context** since you have started using it. What do you think has changed? E.g. change of government/policy around NHS or housing |
| **6. VIGNETTES** |
| I'd like to share a couple of vignettes of data sharing practices to ask your views on these. I'll share the vignette then ask a couple of follow up questions. [*see examples at the end of the guide]* |
| **7. CLOSING** |
| We're reaching the end of the interview. Is there **anything else** you would like to raise?<br>What other information do you think would be useful to us to understand data sharing in your context? *prompts: Who would you recommend we interview? Any documents you can share?* |

**Thank you**

Thank you so much for taking the time to talk to me today – your views are very helpful for our research. Would it be ok to check-in with you if I need to clarify my understanding of that you've shared today?

If you are interested, we will be in touch with the findings from our research. In the meantime, please do get in touch with any questions.

Vignettes to illustrate using shared housing and health data to reduce health inequalities

- In the interview, we would like to learn about your experiences and views of any data sharing you have undertaken or tried or would like to do.
- We appreciate data sharing can seem abstract. We have given below some vignettes adapted from data sharing innovations that colleagues have shared so far already, to give some tangible examples of different types of data sharing.
- We will use 1-2 of these examples to stimulate discussion with you.
- However, you don't need to focus solely on these examples, we would like to hear about the work you have done or would like to do in this area.

**Vignette 1**

**Individual/household level: Sharing NHS data with Local Authority data systems (NB could be anonymised or identifiable)**

Example: Living in cold and damp homes can contribute both to the onset and worsening of physical health conditions, including respiratory and cardiovascular conditions.

A count, from NHS data, on the number of council home residents' health vulnerabilities, e.g. respiratory illness, has been extracted from clinical records. This data has been shared with the council's housing department. So far, it has been used for an audit to compare the repair times for people reporting damp/mould problems amongst those with and without health vulnerabilities. It could also be used for health risk stratification to prioritise housing strategies, e.g. checking ventilation, boiler replacement, help with heating bills, installing remote sensors to monitor temperature/damp levels.

**Vignette 2**

**Individual/household level: Sharing Local Authority data to NHS data systems (NB could be anonymised or identifiable)**

Example: Using rent arrears as a potential priority to target stop smoking outreach services.

Adults living in social housing are more likely to smoke than those living in other housing types and targeted interventions are needed to reduce inequalities in smoking by housing tenure (Smoking in social housing among adults in England, 2015–2020: a nationally representative survey)

The financial benefits of stopping smoking may incentivise residents in rent arrears to quit smoking, if they are made aware of these and signposted to support to do so.

Individual/household level data on rent arrears and smoking status could be used to target stop smoking outreach services or support housing officers to use opportunities arising during routine contacts with residents to support residents to stop smoking.

**Vignette 3**

**Population-level: presentation of aggregate data from multiple sectors on same dashboard**

Children living in overcrowded conditions have an increased risk of developing behavioural issues such as aggression or hyperactivity, increased school absences, and delayed cognitive development. (Building Health Equity: The Role of the Property Sector in Improving Health)

A dashboard integrating cross sector local data on overcrowding, educational attainment and mental health of children and young people living in temporary accommodation supports local authorities and health partners to better understand the local situation and how to respond.

**Vignette 4**

**Individual/household level: Residents provide data on their own health conditions to housing services or housing conditions to health services**

The Social Housing Act places an obligation on providers to have robust information about their residents. Information on residents' health conditions or mobility constraints can be useful for housing teams to identify those most at risk in emergencies (e.g. wheelchair users when lifts are broken or to inform fire evacuation procedures). Housing providers are exploring multiple ways to collect and update this information from residents, including emailed surveys, opportunistic collection when contacted about another matter.

Conversely, information on patients' housing conditions can be useful for primary care teams to inform management of long-term conditions. Some primary care services are asking all patients to provide this information.

**Vignette 5**

**Individual-level data shared between professionals in context of joint working (e.g. referrals or MDT meetings)**

Joint working can take multiple forms, but would typically involve sharing data, e.g. at MDT meetings in referral pathways between health and housing services.

**Indoor Air Quality**

NICE Indoor Air Quality guidance (2020) recommends joint working between local authority departments, with local health and social care providers, and with voluntary, community and social enterprise organisations and other organisations with interest in indoor air quality.

Examples of joint working have been shared for the purposes of:

- preventing delayed hospital discharge due to housing-related reasons or
- implementing evidence-based care for asthma patients living in homes with damp/mould.

**Supporting people with complex mental health conditions**

Joint working between housing, health and wider partners could provide more holistic support to residents with complex mental health conditions and support tenancy sustainment. This may include preventing eviction e.g. due to self-neglect or hoarding, or after being sectioned in hospital.

<u>Questions on data sharing</u>

- At what point would it be necessary/appropriate to ask permission to access data? And from whom? Resident? Service provider?
- How might this information be used? (by clinicians or housing departments)
    - What is the currency and format of data needed to inform action? e.g. respiratory disease prevalence, frequency/type of healthcare use for respiratory symptoms, extent (or risk) of damp/mould, other family, clinical, environmental and social determinants of health data that could affect health and wellbeing or access/receptiveness to interventions
    - What else is needed to make the information useful? E.g. training/briefings on respiratory disease? Integration of information into the repairs pathway /workflow
- What **barriers or challenges** to sharing such data do you envisage (e.g. fears of performance management, compromising patient/resident privacy, breaches of information governance, difficulties in matching or standardising data across organisations)
- Possible unintended consequences, e.g. flagging damp/mould & health problem could increase risk of eviction?
- Envisaged effects on inequalities – positive and negative, e.g.
    - Differences in acceptability of data sharing for different groups
    - Potential for widening inequalities?
